# Development of Moore Swab and Ultrafiltration Concentration and Detection Methods for *Salmonella* Typhi and *Salmonella* Paratyphi A in Wastewater and Application in Kolkata, India and Dhaka, Bangladesh

**DOI:** 10.1101/2021.03.20.21254025

**Authors:** Pengbo Liu, Makoto Ibaraki, Renuka Kapoor, Nuhu Amin, Abhishek Das, Md. Rana Miah, Asish K Mukhopadhyay, Mahbubur Rahman, Shanta Dutta, Christine L. Moe

## Abstract

Enteric fever is a severe systemic infection caused by *Salmonella enterica* serovar Typhi (ST) and *Salmonella enterica* serovar Paratyphi A (SPA). Detection of ST and SPA in wastewater can be used as a surveillance strategy to determine burden of infection and identify priority areas for water, sanitation, and hygiene interventions and vaccination campaigns. However, sensitive and specific detection of ST and SPA in environmental samples has been challenging. In this study, we developed and validated two methods for concentrating and detecting ST/SPA from wastewater: the Moore swab trap method for qualitative results, and ultrafiltration (UF) for sensitive quantitative detection, coupled with qPCR. We then applied these methods for ST and SPA wastewater surveillance in Kolkata, India and Dhaka, Bangladesh, two enteric fever endemic areas. The qPCR assays had a limit of detection of 17 equivalent genome copies (EGC) for ST and 25 EGC for SPA with good reproducibility. In seeded trials, the Moore swab method had a limit of detection of approximately 0.05-0.005 cfu/mL for both ST and SPA. In 53 Moore swab samples collected from three Kolkata pumping stations between September 2019 to March 2020, ST was detected in 69.8% and SPA was detected in 20.8%. Analysis of sewage samples seeded with known amount of ST and SPA and concentrated via the UF method, followed by polyethylene glycol precipitation and qPCR detection demonstrated that UF can effectively recover approximately 8 log_10_ cfu, 5 log_10_ cfu, and 3 log_10_ cfu of seeded ST and SPA in 5 L, 10 L, and 20 L of wastewater. Using the UF method in Dhaka, ST was detected in 26.7% (8/30) of 20 L drain samples with a range of 0.11-2.10 log_10_ EGC per 100 mL and 100% (4/4) of 20 L canal samples with a range of 1.02 - 2.02 log_10_ EGC per 100 mL. These results indicate that the Moore swab and UF methods provide sensitive presence/absence and quantitative detection of ST/SPA in wastewater samples, and these two methods can be used jointly or separately for *Salmonella* Typhi environmental surveillance.

## Introduction

Typhoid and Paratyphoid fevers are leading causes of severe febrile disease in low-income countries with poor access to safe water, food, and sanitation.^1^ The etiologic agents, *S*. Typhi (ST) and *S*. Paratyphi A (SPA), are human-specific pathogens transmitted through consumption of food and water contaminated by feces of an acutely or chronically infected person.^2^ Previous studies have shown that food and water contaminated with human feces are associated with typhoid and paratyphoid outbreaks.^3, 4^ In endemic urban settings, co-location of sewer pipes and poorly maintained water supply pipes facilitate cross-contamination and transmission of ST through the water system. Typhoid fever outbreaks^4, 5^ have been associated with contaminated piped water and sewage-irrigated produce, emphasizing the importance of studying these pathways of disease transmission.

Detection of ST and SPA in drinking water, irrigation water, and environmental waters has usually been attempted in association with outbreak investigations^6, 7^ and assessing risk of waterborne typhoid fever transmission in endemic settings.^8^ Recently, we and other investigators have proposed detection of ST and SPA in wastewater from known catchment populations as a strategy to determine the burden of typhoid and paratyphoid fever in areas where clinic-based surveillance has limited sensitivity or is not feasible.^9, 10^ However, environmental surveillance for typhoid and paratyphoid fever requires sensitive and specific methods to detect ST/SPA in wastewater, including samples where low pathogen concentrations are expected. It is usually necessary to concentrate ST/SPA in environmental samples in order to increase the sensitivity of detection. A wide range of methods have been used to concentrate bacterial pathogens from a variety of environmental waters.^10^ Among these methods, Moore swabs and ultrafiltration (UF) are two important and distinct concentration methods that have been shown to be effective in recovering ST from water and wastewater.^11^ The Moore swab was first introduced for *Salmonella* detection from sewage in 1948 in England during a paratyphoid epidemic.^6^ Subsequently, this method has been successfully used to isolate *Vibrio Cholerae*^12^, poliovirus^13^ and *Burkholderia pseudomallei*^14^ from sewage. In contrast to the Moore swab method that only shows the presence or absence of a target pathogen, UF is a quantitative method that can simultaneously concentrate multiple pathogens from large volumes of water or wastewater,^15-17^ and we recently reported the application of this method to detect *S*. Typhi in wastewater samples.^18^ The Moore swab method offers several advantages over UF, specifically, Moore swabs are inexpensive, simple to use, and do not require collecting and transporting large volume samples of water or wastewater. However, UF can provide quantitative results with greater sensitivity which are valuable for microbial risk assessment or estimating infection prevalence.^18^

The ability to detect and quantify ST/SPA in the environment is critical for monitoring and controlling transmission particularly in urban settings in low- and middle-income countries. Historically, culture-based isolation and identification methods are considered the gold standard because infectious bacteria can be detected. However, culture of *S*. Typhi from environmental samples is challenging.^19^ Alternatively, PCR and qPCR technologies have been widely used to rapidly detect and quantify ST/SPA due to higher sensitivity and specificity and short turnaround time. Limitations of molecular detection methods include the inability to distinguish between infectious and non-infectious ST/SPA cells, and the presence of PCR inhibitors in environmental matrices in samples that can potentially lead to an underestimation of the target nucleic acid or false negative results.

The overall goal of this study was to develop a qualitative and a quantitative method for ST and SPA detection in wastewater and apply these methods to detect ST and SPA in two typhoid fever endemic areas. The specific goals were to: (1) develop real-time qPCR methods with standard curves for detecting and quantifying ST and SPA DNA in sewage; (2) develop and validate Moore swab (qualitative) and UF concentration methods (quantitative); (3) apply the Moore swab method to detect ST/SPA in wastewater at three pumping stations in Kolkata, India; and 4) utilize the UF method to detect ST in drain and canal water samples in Dhaka, Bangladesh.

## Materials and Methods

### Study sites and environmental sample collection

Wastewater collected from the WaterHub water reclamation facility on the Emory University campus between April 2019 and July 2020 were used for the Moore swab and UF methods development and validation studies in Atlanta. From April to October 2019, drain and canal water samples were collected in Mirpur, Dhaka, Bangladesh, a densely populated low-income area where residents live in compounds containing multiple families and the incidence of ST and SPA is high.^18, 20^ Large volume (20 L) samples were collected and concentrated by UF as described below. In Kolkata, India, Moore swabs were placed in three municipal sewage pumping stations between September 24, 2019 and March 19, 2020: 1) Palmer’s Bridge station served Ward 55 with approximately 32,254 people; 2) Ambedkar station served Wards 66 and 108 with approximately 162,801 people; and 3) Topsia station served Wards 59 and 48 with approximately 90,698 people during the study period.

### ST and SPA growth conditions

ST (ATCC 19430) and SPA (ATCC 9150) cultures were prepared by overnight growth in LB (Luria-Bertani) broth at 37°C under shaking conditions to obtain a final concentration of 10^8^ cells/mL, followed by 10-fold serial dilutions in 1× PBS (0.01 M, pH 7.4). One milliliter aliquots of the diluted culture containing 10^8^ to 100 cells were used for seeding experiments.

### ST and SPA DNA standard development

ST and SPA DNA were extracted from the overnight fresh culture, and the DNA concentration was measured using NanoDrop spectrophotometer (Thermo Fisher Scientific, Waltham, MA USA). The concentrations and equivalent genome copies (EGC) of the ST and SPA DNA were determined from the concentration (ng/µL), the lengths of fragments, the Avogadro constant, and the average weight of double-stranded base pairs (equation 1).

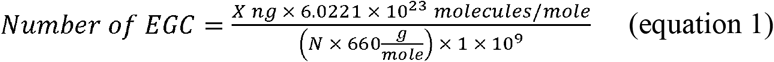

Where: *X* = amount of amplicon (ng)

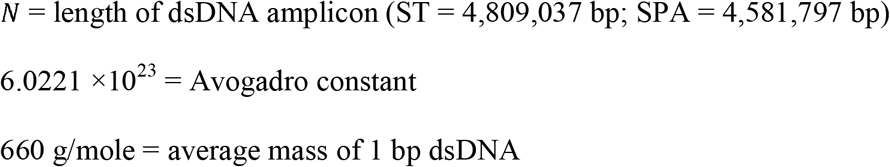

The known EGCs for each standard were serially diluted, and a standard curve was incorporated in each real-time PCR assay. Ten-fold serial dilution of the standards was used to estimate the numbers of genome copies of the target pathogens in samples.

### Ultrafiltration method validation

Large volume wastewater samples (5-20 L) were seeded with known amounts of ST and SPA (either 8 log_10_ CFU, 5 log_10_ CFU, or 3 log_10_ CFU), and then each sample was concentrated by hollow fiber ultrafiltration (Figure 1). The entire amount of the seeded wastewater sample was circulated through Polynephron^™^ Synthetic Hollow-Fiber Dialyzer (NIPRO Medical United States, Bridgewater, USA) using a peristaltic pump to achieve approximately 100 mL of retentate (concentrated sample) which was collected in a 500 mL bottle. Ultrafilter elution was then performed using 500 mL of PBS with 0.01% Tween 80, 0.01% sodium polyphosphate and 0.001% Antifoam Y-30 emulsion. The elution solution was recirculated for 5 mins, and the final eluate was merged into the concentrated sample. After elution, the ultrafilter was backwashed using 250 mL of PBS with 0.5% Tween 80, 0.01% sodium polyphosphate and 0.001% Antifoam Y-30 emulsion. The backwash fraction was added to the previous mixture of the concentrate and the eluate.

**Figure 1.**
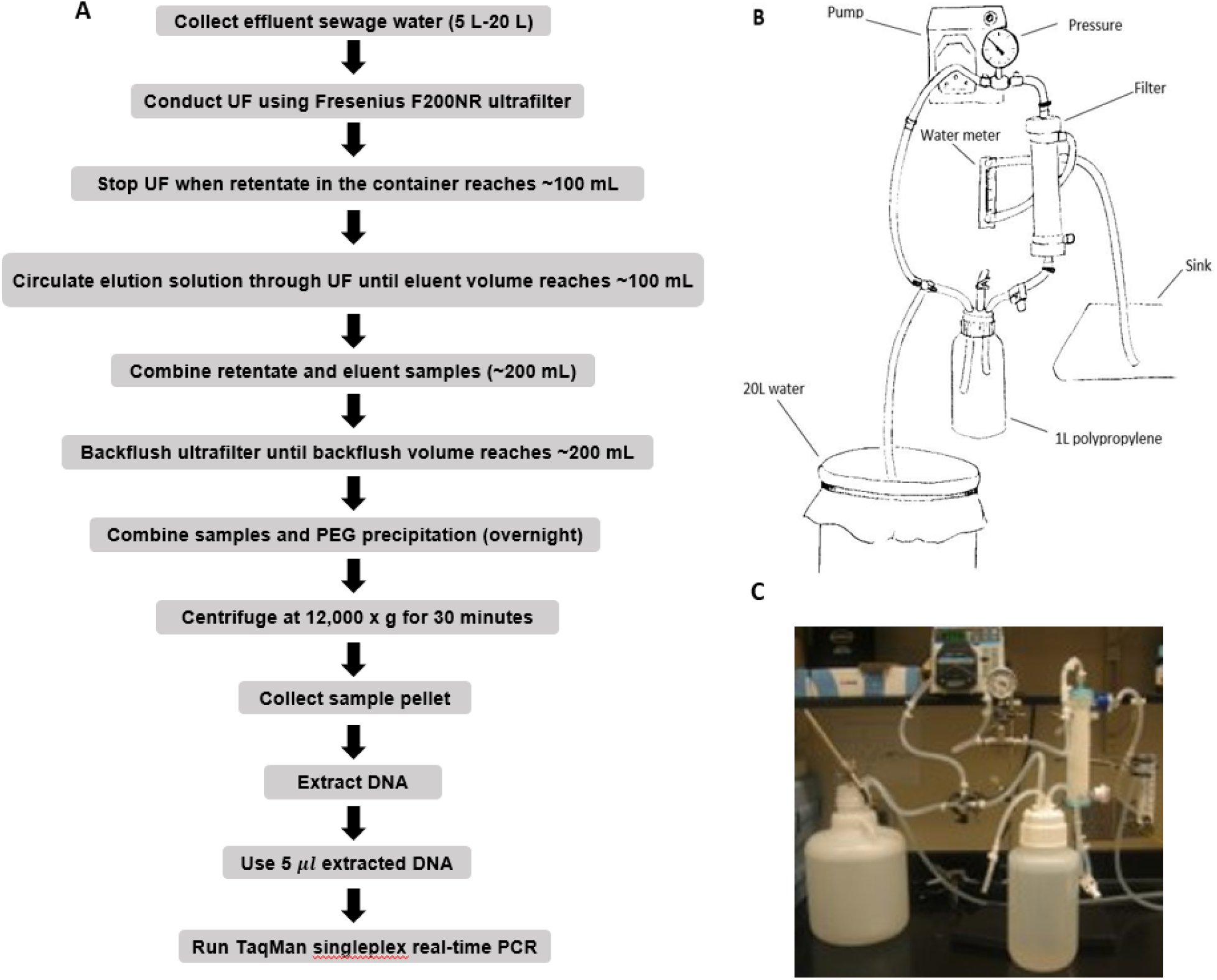
Ultrafiltration method for recovering seeded *S*. Typhi and *S*. Paratyphi A from wastewater samples. 1A. Flow diagram showing procedures for ST/SPA concentration and detection from seeded wastewater or field samples. 1B. Schematic of hollow-fiber ultrafiltration setup. 1C. Hollow-fiber ultrafiltration laboratory setup.

### ST and SPA concentration using polyethylene glycol (PEG)

ST and SPA were precipitated from the merged retentate/eluate/backflush samples by adding 12% polyethylene glycol 8000 (Sigma, St. Louse, MO, USA), 0.9 mol sodium chloride, 1% bovine serum albumin (Sigma) and incubated and stirred overnight at 4°C. After centrifugation at 12,000 g for 60 min, the pellet was suspended in 1 mL of InhibitEX buffer provided by the QIAamp Fast DNA Stool Mini Kit (Qiagen, Valencia, CA, USA) prior to DNA extraction.

### Moore swab method validation

Moore swabs (Figure 2B) were made by cutting pieces of cotton gauze to approximately 120 cm long × 15 cm wide and firmly tying the center with fishing line (W.C. Bradley/Zebco Holdings Inc., Tulsa OK). The swabs were sterilized before use by autoclaving. A Moore swab was placed in a plastic container filled with 2-20 L sewage collected from the Emory WaterHub, with the end of the fishing line attached to the outside of the container (Figure 2C). Sewage samples were seeded with either 50 cfu/mL, 20 cfu/mL, 10 cfu/mL, 5 cfu/mL, 0.05 cfu/mL, or 0.005 cfu/mL of ST and SPA cells. The swab was submerged in the sewage to trap the ST and SPA cells while stirred continuously with an overhead spatula for 24 hours as shown in Figure 2C. The swabs were then transferred to 450 mL of universal pre-enrichment broth (EPA, Standard Analytical Protocol for *Salmonella* Typhi in Drinking Water) and incubated at 37°C for 24 hrs with shaking. Then, a 20 mL volume of the pre-enrichment broth was filtered through a 0.45 µm filter and subjected to DNA extraction as described below after the addition of 1 mL of InhibitEX buffer (Figure 2A).

**Figure 2.**
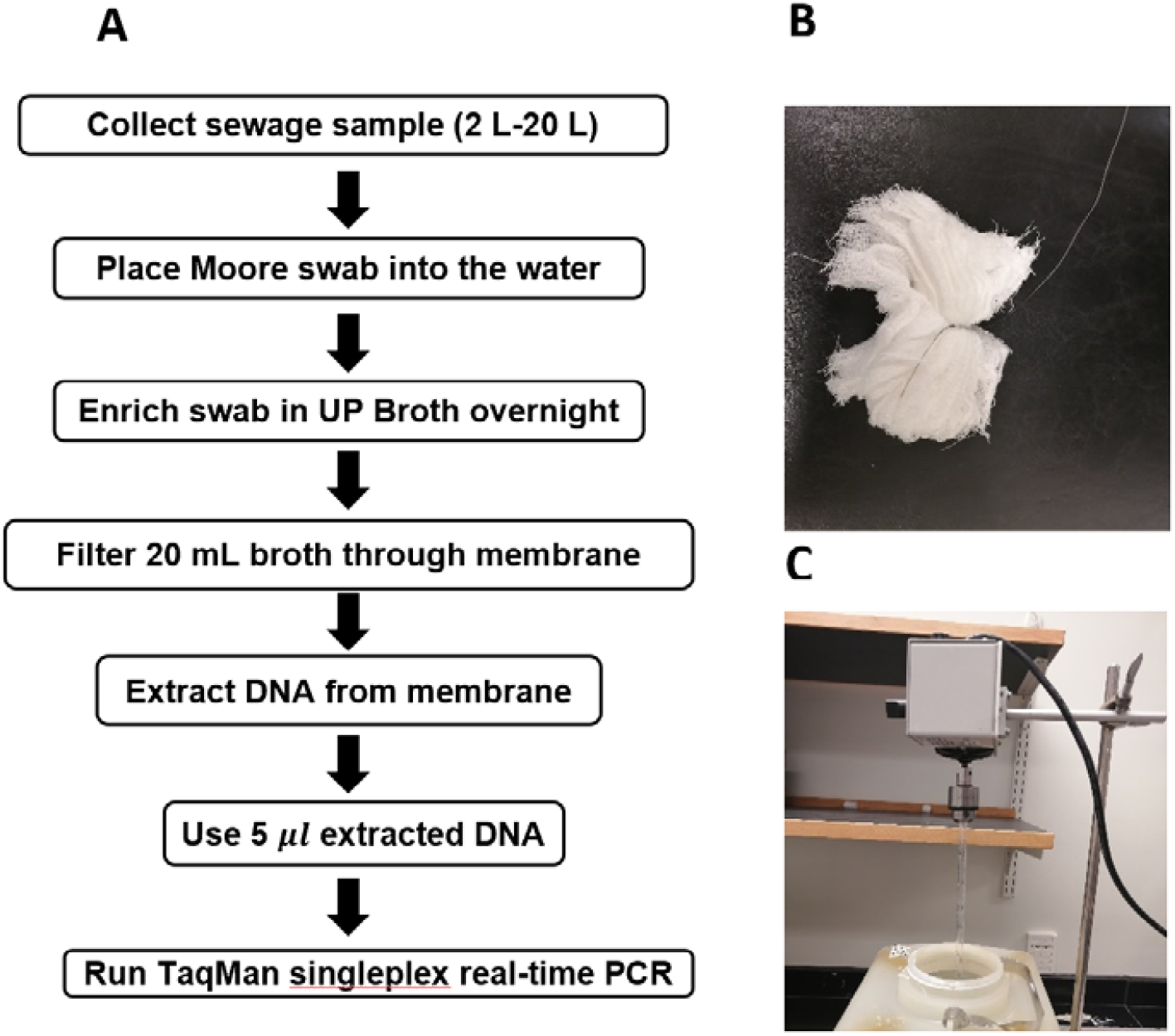
Moore Swab method for recovering *S*. Typhi and *S*. Paratyphi A from seeded sewage samples. 2A. Flowchart for the Moore swab method validation experiments. 2B. Moore swab. 2C. Moore swab method laboratory setup for seeded sewage samples.

### ST/SPA DNA extraction

For each sample or blank control, 1 mL of the final sample suspension, mixed with the InhibitEX buffer, was transferred into a 2 mL microcentrifuge tube. For membrane-filtered samples and controls, microcentrifuge tubes with the filter and 1 mL of InhibitEX buffer were used. After one minute of vortexing, the solution was centrifuged at maximum speed for 2-3 minutes, and the suspension was incubated at 95°C for 5 minutes, followed by full speed microcentrifugation for 15 seconds to pellet the sample particles. Subsequently, 600 µl of the supernatant was transferred into a 2 ml microcentrifuge tube containing 25 µL proteinase K, and then 600 µL of buffer AL was added. After vortexing for 15 seconds, the sample was incubated at 70°C for 10 minutes, and 600 µL of 100% ethanol was added to the lysate. A 600 µL volume of lysate was applied to a single QIAamp Mini column (Qiagen, Cat. # 51604, Hilden, Germany), and the column was centrifuged at full speed for 1 minute. This procedure was repeated two additional times with the same column in order to use up all of the lysate. Finally, TNA was eluted from the column with 150 µL of the supplied elution buffer. The extracted TNA samples were aliquoted and stored at −80°C until analyzed by qPCR.

### TaqMan singleplex real□time PCR for detection of ST and SPA DNA

Quantitative singleplex real-time PCR assays for ST and SPA detection were performed using a Bio-Rad CFX96 thermal cycler (Bio-Rad Laboratories, Inc, Berkeley, CA). The SPA PCR primers and probe described by Karkey et al^21^ were used for these laboratory assay evaluation experiments. For SPA detection in Moore swab samples in Kolkata, newly-developed primers and probe targeting the intergenic region SSPAI (between SSPA1723a and SSPA1724) were used, and their sequences are as follows; SPA2RK_F: 5’-ACCATCCGCAGGACAAATC-3’; SPA2RK_R: 5’-GGGAGATTACTGATGGAGAGATTAC-3’; SPA2RK_Probe: 5’-Cy5-AGAG TGCAAGTGGAGTGCCTCAAA-BHQ2-3’.

For ST detection, two sets of primers/probe were used: 1) the primers/probe described by Karkey et al.^21^ for the samples concentrated by UF in Dhaka, and 2) a newly-designed ST primers/probe set for the method validation work in Dr. Moe’s lab and the Moore swab samples in Kolkata. The newly-designed ST primers/probe targeted the ST stgA gene (codes for putative fimbrial subunit protein) and the sequences are as follows: ST2RK_F (forward): 5’-TATCGGCAACCCTGCTAATG-3’; ST2RK_R (reverse) 5’-TATCCGCGCGGTTGTAAAT-3’; ST2RK_Probe 5’ FAM-CCATTACAG CATCTGGCGTAGCGA-BHQ1-3’. PCR reactions were performed in 25 µl volumes consisting of 2 × Bio-Rad iQ power mix buffer (Bio-Rad, Hercules, CA) including 200 µM dNTPs, 12 mM MgCl2, 1 U iTaq DNA polymerase, 400 nM of each primer, 200 nM of each probe, and 5 µL of template TNA or negative control. The amplification procedure consisted of a preliminary denaturation step at 95°C for 3 minutes and 45 cycles of 95°C for 30 seconds, 60°C for 30 seconds, and 72°C for 30 seconds. Fluorescence was collected at the annealing steps during the last cycles. To quantitatively detect equivalent genome copies of each pathogen in the samples, a ten-fold serial dilution of standards was added to each set of assay plates.

### Data analysis

ST and SPA concentrations in each sample were estimated by interpolation of the mean Ct values from duplicate wells to the standard curve, generated by 10-fold series diluted ST and SPA DNA standards, and the wastewater concentration factor. The quantification was expressed as log_10_ EGC per 100 mL wastewater. Experiments evaluating the limit of detection of the Moore swab and UF concentration method were repeated three times for each volume and reported seeding level. Experiments for assessing ST and SPA recovery efficiencies were also repeated three times for each volume and seeding level. The number of equivalent genome copies in each test sample (*N*_sample_) was estimated based on the linear regression of log_10_ (*N*_sample_) versus Ct values using equation 2. The amplification efficiency was estimated in each real-time PCR run using the slope m of the standard curve (equation 3).

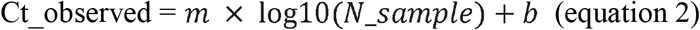

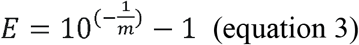

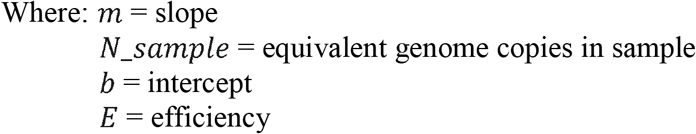

## Results

1. Performance of ST and SPA qPCR Assays and DNA Standards Two singleplex TaqMan real-time qPCR assays were evaluated for the detection of ST and SPA species using primers/probe previously described by Kaykey et al.^21^ To evaluate the analytical sensitivity of the two qPCR assays, DNA from ST strain (ATCC 19430) and SPA strain (ATCC 9150), ranging from 1.7 × 10^4^ to 17 EGCs for ST, and 2.5 × 10^4^ to 25 EGC for SPA per reaction, were tested. Quantitative detection was achieved in the range of template DNA concentrations and the Ct values with R^2^ (coefficient of determination) between 0.98-1.00 for ST and 0.97-0.99 for SPA (Table 1 and Figure 3). The limit of detection was 17 EGC per reaction for ST and 25 EGC per reaction for SPA. The qPCR efficiency was between 76.3-127.9% for ST and 81.9-99.0% for SPA. The assay coefficient of variation (CV) was calculated by measuring the variation in the Ct values separately for the high (4-log_10_) and low concentrations (1-log_10_) in the range of standard concentrations in three replicate experiments on different days. The inter-assay CVs of the high and low ST concentrations from three replicate experiments ranged from 1.81-2.89%, respectively, whereas the inter-assay CVs of the high and low SPA concentrations were 1.59% and 2.08%, respectively (Table 1). These results demonstrated that both ST and SPA qPCR assays were sensitive and reproducible.

**Table 1.** Performance of S. Typhi and S. Paratyphi qPCR Assays^†^ and DNA Standards.

| Pathogen | Strain | qPCR<br>Replicates | Efficiency<br>(%) | R<sup>2</sup> | LOD<sup>*</sup><br>(EGC) | High Concen.<br>CV<sup>#</sup> (%) | Low Concen.<br>CV (%) |
| --- | --- | --- | --- | --- | --- | --- | --- |
| ST | ATCC<br>19430 | 3 | 76.3-127.9 | 0.98-1.00 | 17 | 1.81 | 2.89 |
| SPA | ATCC<br>9150 | 3 | 81.9-99.0 | 0.97-0.99 | 25 | 1.59 | 2.08 |

<sup>†</sup>Using primers/probes described by Kaykey et al<sup>21</sup>
<sup>\*</sup>Limit of detection
<sup>#</sup>Coefficient of variation $CV = \frac{\sigma}{\mu}$ $\sigma$ : standard deviation; $\mu$ : mean

**Figure 3.**
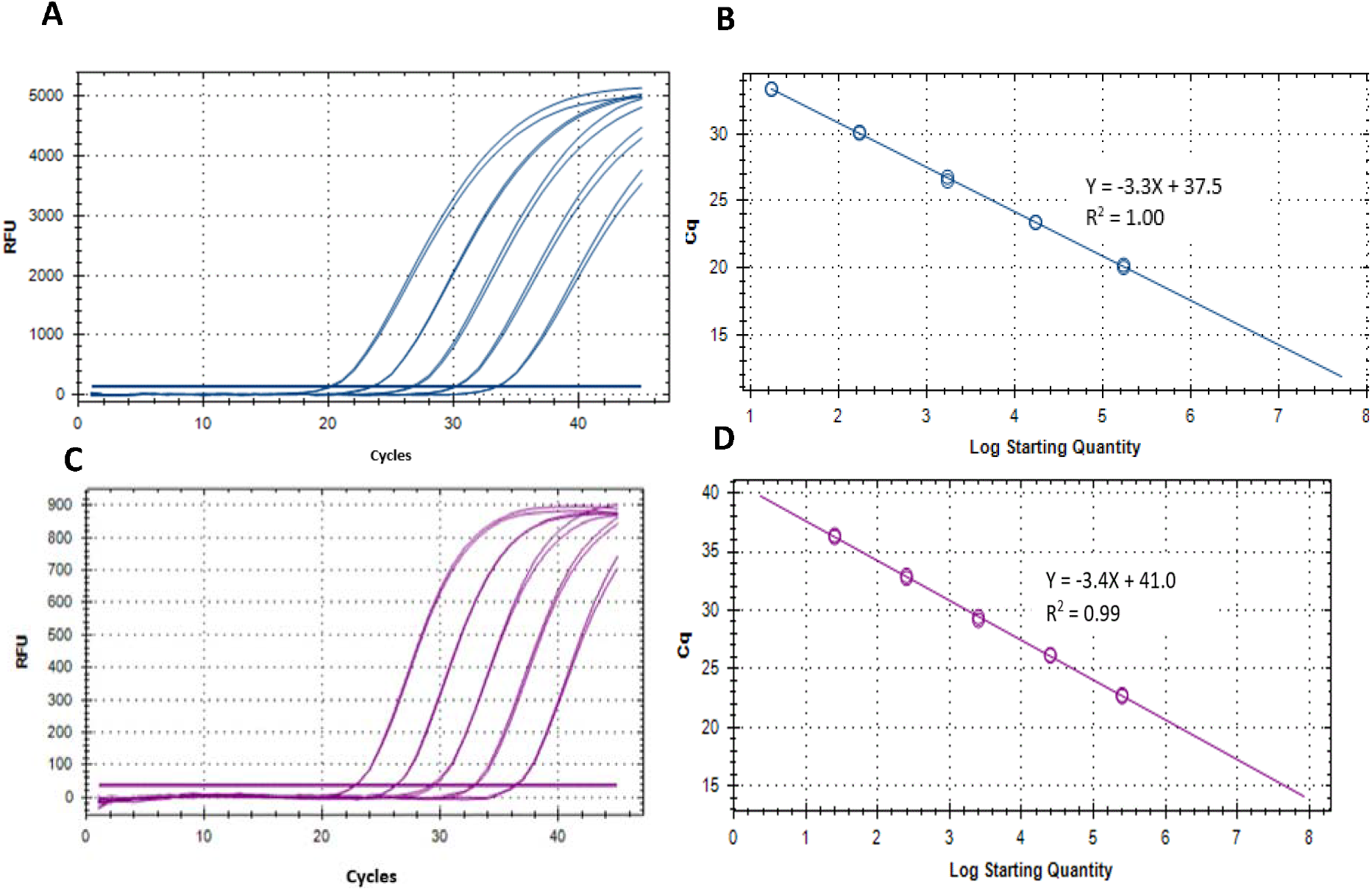
Performance of *S*. Typhi and *S*. Paratyphi A qPCR assays described by Karkey et al^21^ and DNA standards. A) Amplification of ten-fold serially-diluted *S*. Typhi DNA standard. B) Standard curve of *S*. Typhi DNA. C) Amplification of ten-fold serially diluted *S*. Paratyphi A DNA standard. D) Standard curve of *S*. Paratyphi A DNA.
2. Limit of Detection of Moore Swabs for recovering seeded ST and SPA in different volumes of sewage samples To determine the applicability and limit of detection of the Moore swab method and qPCR for the detection of ST and SPA in wastewater, samples of wastewater were collected from the Emory WaterHub and seeded with known concentrations (50 cfu/mL, 20 cfu/mL, 10 cfu/mL, 5 cfu/mL, 0.05 cfu/mL, 0.005 cfu/mL) of ST and SPA. When both ST and SPA were seeded at high concentrations (50 cfu/mL, 20 cfu/mL, 10 cfu/mL, and 5 cfu/mL), all replicate experiments showed positive qPCR results. When the seeding level was reduced to 0.05 cfu/ml, ST was detected in all three replicate experiments, but SPA was only detected in two of the three replicate experiments. When the seeding level was further reduced to 0.005 cfu/mL, both SPA and ST were detected in all three replicate experiments (Table 2). Lower seeding concentrations of ST and SPA were not examined due to the high Ct values (38-40) that were observed at the 0.005 cfu/mL seeding level. These results indicate the limit of detection of both ST and SPA using the Moore swab method was approximately 0.05-0.005 cfu/mL.

**Table 2.**
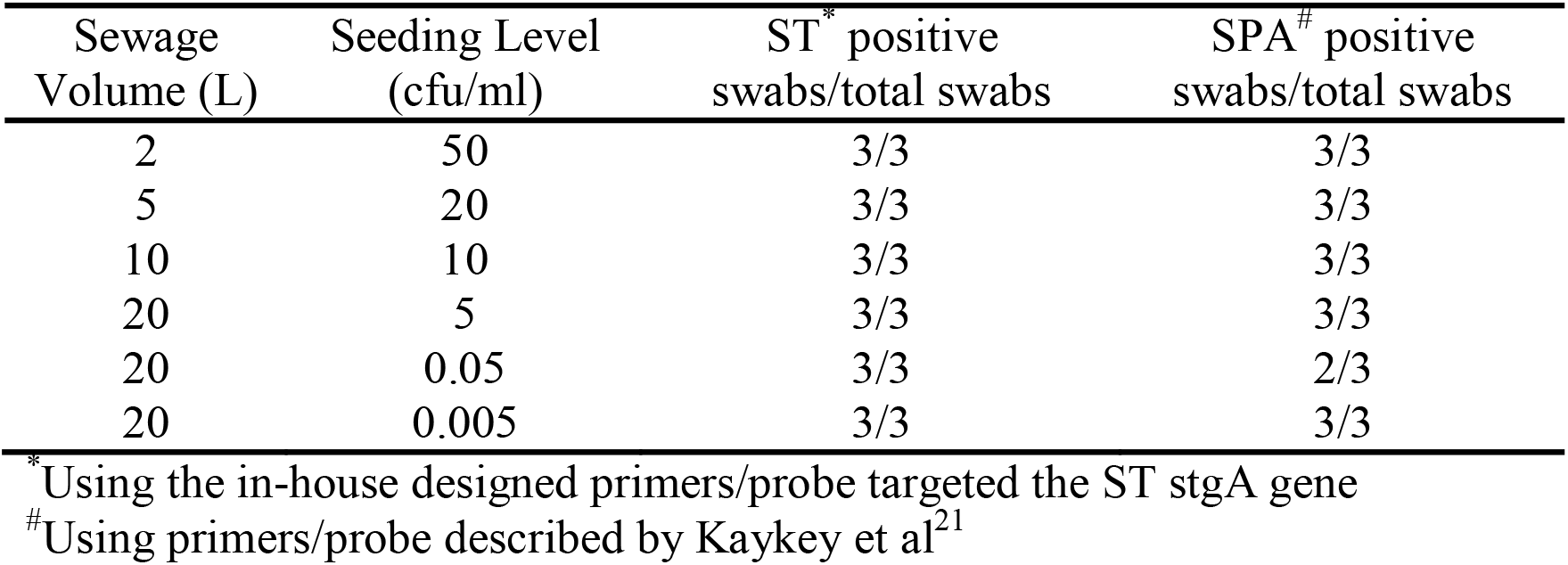
Limit of Detection of Moore Swab method for recovering ST and SPA seeded into different volumes of sewage samples.
3. Quantification of ST and SPA recovered from seeded sewage samples using UF 3a. Quantification of ST and SPA recovered from different seeding levels in 20 L sewage samples Twenty-liter sewage samples seeded with known amounts of ST and SPA were concentrated by UF, followed by PEG precipitation and qPCR detection. After seeding an average of 9.13 log_10_ cfu ST in 20 L of sewage and concentrating the sample using UF, the ST qPCR assay detected a mean of 6.84 log_10_ EGC ST in three replicate experiments. When an average of 5.71 log_10_ cfu ST was seeded into 20 L samples in three experiments, 3.20 log_10_ EGC ST was recovered. Seeding 20 L wastewater samples with a mean total of 3.49 log_10_ cfu ST resulted in the recovery of an average 2.81 log_10_ EGC ST (Figure 4A). Similarly, seeding an average total of 7.94 log_10_ cfu SPA led to a mean total recovery of 5.80 log_10_ EGC SPA. When an average total of 5.0 log_10_ cfu SPA was seeded, an average total of 3.62 log_10_ EGC SPA was recovered. Seeding with a mean total of 4.07 log_10_ cfu SPA resulted a total of 3.32 log_10_ EGC SPA recovery (Figure 4B). These results indicate that UF followed by PEG concentration can effectively recover different concentrations of ST and SPA seeded in large volumes of sewage, but there is about a 2 log_10_ loss of the target bacteria during the concentration, TNA extraction, and qPCR process.

**Figure 4.**
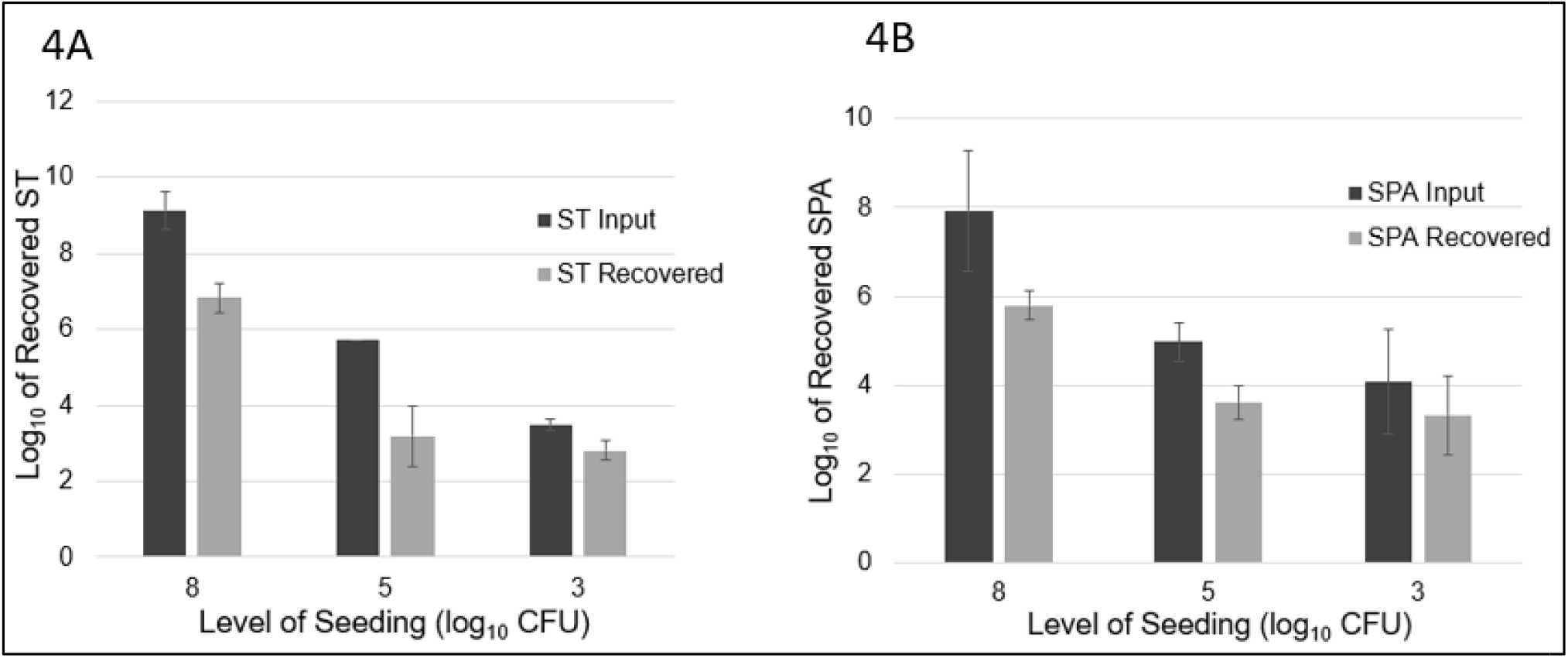
ST and SPA mean recovery at different seeding levels (approximately 8 log_10_ cfu, 5 log_10_ cfu, and 3 log_10_ cfu) in 20 L of sewage using the ultrafiltration method in three replicate experiments. The experiments were validated using *Salmonella* strains from ATCC (ST: 19430; SPA:9150) tested by primers/probes described by Kaykey et al.^21^ Error bars represent standard deviation in three replicate experiments. 4A. Black bars represent the mean input of ST seeded into the 20 L sewage samples, and gray bars represent the mean recovered ST. 4B. Black bars represent the mean amount of SPA seeded into the 20 L sewage samples, and gray bars represent the mean amount of SPA detected by qPCR. 3b. Quantification of ST and SPA recovered from different volume of seeded sewage To examine if sample volume affects ST and SPA recovery using UF, an average total of 9.23 log_10_, 8.54 log_10_, and 9.13 log_10_ cfu ST were seeded into 5 L, 10 L, and 20 L sewage samples, respectively. Mean totals of 7.57 log_10_, 6.13 log_10_, and 6.84 log_10_ EGC ST were recovered (Figure 5A), respectively. Similarly, when an average total of 8.73 log_10_, 8.74 log_10_, and 7.94 log_10_ cfu SPA was seeded into 5 L, 10 L, and 20 L of sewage water, respectively, a mean total of 6.82 log_10_, 5.97 log_10_, and 5.70 log_10_ EGC SPA was recovered, respectively (Figure 5B). These results indicate that sample volumes in the range of 5-20 liters did not affect the efficiency of ST and SPA recovery by UF.

**Figure 5.**
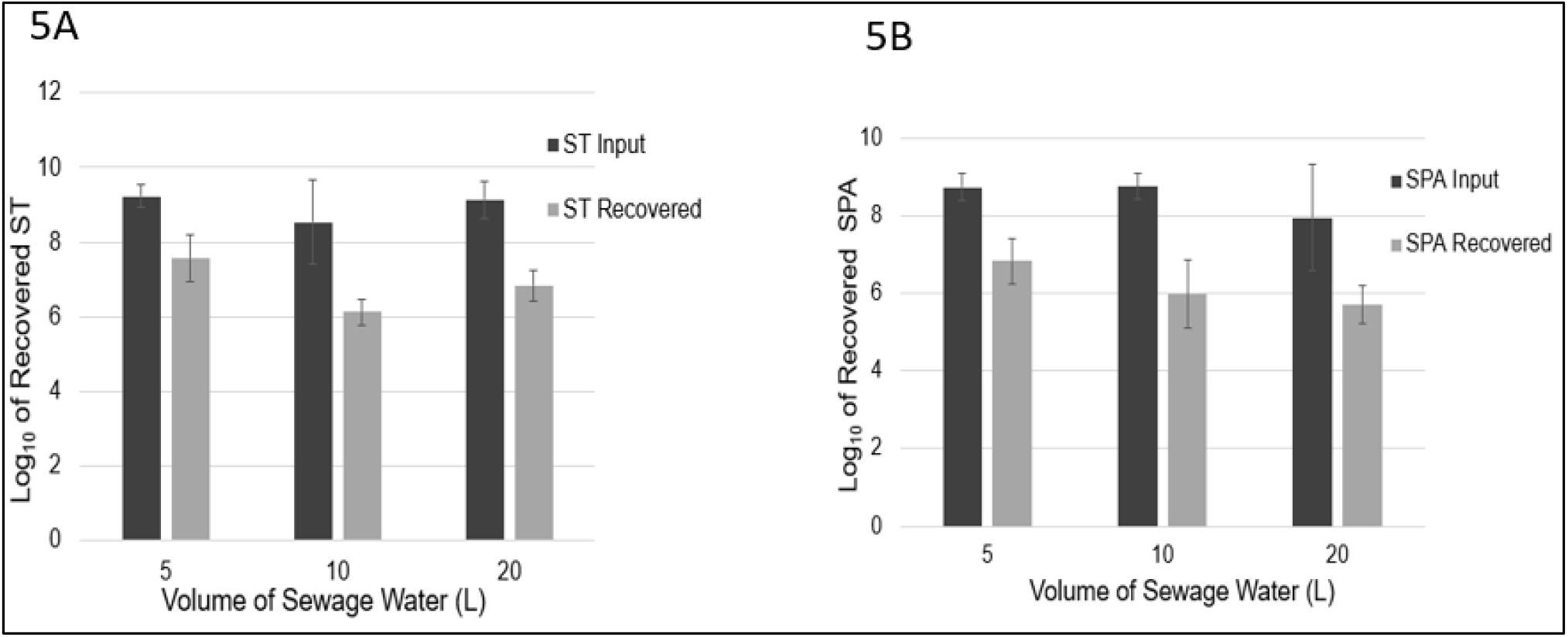
ST and SPA mean recovery in different volumes (5 L, 10 L, and 20 L) of sewage samples using the ultrafiltration method in three replicate experiments. Error bars represent standard deviation. The experiments were validated using *Salmonella* strains from ATCC (ST: 19430; SPA:9150) tested by primers/probes described by Kaykey et al.^21^ 5A. Black bars represent the mean input ST seeded into 5 L, 10 L and 20 L samples in three replicate experiments, and gray bars represent the mean recovered detected by qPCR. 5B. Black bars represent the mean SPA seeded, and gray bars represent the mean recovered SPA detected by qPCR.
4. ST and SPA detection in 20 L Environmental Water Samples concentrated by UF in Dhaka, Bangladesh In the study neighborhood in Dhaka, wastewater from toilets shared by multiple households discharges directly into open drains and canals adjacent to the streets. ST was detected in 26.7% (8/30) of drain samples and 100% (4/4) of canal samples that were concentrated by UF. Estimated concentrations of positive ST detected in drain samples ranged from 0.23 to 1.26 log_10_ EGC/100 mL with a mean of 0.82 log_10_ EGC/100 mL. In the canal samples, the estimated ST concentrations ranged from 0.71 to 2.14 log_10_ EGC/100 mL (mean 1.43 log_10_ EGC/100 mL) (Table 3). These results indicate that ultrafiltration is an effective quantitative method for ST detection in wastewater samples.

**Table 3.** ST Detection using the primers/probe described by Karkey et al ^21^ in 20 L of Environmental Water Samples Concentrated by Ultrafiltration in Dhaka, Bangladesh, April to October 2019.
5. ST and SPA Detection in Moore Swab Samples from Three Pumping Stations in Kolkata, India Between September 24, 2019 to March 19, 2020, 14 Moore swabs were placed on a weekly basis at the Palmer Bridge pumping station, and ST was detected in 10 (71.4%) and SPA was detected in 4 (28.6%) of these swabs. Weekly Moore swabs were also placed in the Ambedkar Bridge pumping station and Topsia pumping station. ST and SPA were detected in 63.2% (12/19) and 31.6% (6/19) of the swabs at the Ambedkar Bridge pumping station, respectively. At the Topsia pumping station, ST and SPA were detected in 75.0% (15/20) and 5.0% (1/20) of the swabs, respectively (Table 4). These results suggest that ST and SPA infections are endemic in the city wards in Kolkata that are served by these pumping stations, and that the Moore swab method is an effective low-cost method for ST and SPA sewage surveillance.

| Sample Type | No. Samples | No. Positive (%) | Mean <sup>*</sup> | 95% CI <sup>#</sup> Low | 95% CI High |
| --- | --- | --- | --- | --- | --- |
| Drain | 30 | 8 (26.7) | 0.82 | 0.23 | 1.26 |
| Canal | 4 | 4 (100.0) | 1.43 | 0.71 | 2.14 |
| Total | 34 | 12 (35.3) | 1.02 | 0.81 | 1.23 |

<sup>\*</sup>log<sub>10</sub> CFUEGC/100 mL
<sup>#</sup>Confidence interval

**Table 4.** ST and SPA Detection Between September 2019 to March 2020 in Moore Swab Samples from three Pumping Stations in Kolkata, India.

| Pumping Station | No. Swab | ST Positive (%) <sup>*</sup> | SPA Positive (%) <sup>#</sup> |
| --- | --- | --- | --- |
| Palmer Bridge | 14 | 10 (71.4) | 4 (28.6) |
| Ambedkar Bridge | 19 | 12 (63.2) | 6 (31.6) |
| Topsia Bridge | 20 | 15 (75.0) | 1 (5.0) |
| Total | 53 | 37 (69.8) | 11 (20.8) |

<sup>\*</sup>Using the in-house designed primers/probe targeted the ST stgA gene
<sup>#</sup>Using primers/probe described by Kaykey et al <sup>21</sup>

## Discussion

Environmental surveillance offers a low-cost, non-invasive, and sensitive strategy to characterize the burden of infection in specific populations for pathogens that may be challenging to diagnose through collection and analyses of clinical specimens – such as poliomyelitis, typhoid fever, and recently, COVID-19.^22-25^ The goal of this study was to develop and validate sensitive and specific methods for concentration and PCR detection of ST and SPA in wastewater for typhoid environmental surveillance and also in other environmental samples that could serve as vehicles of ST and SPA transmission. We report here that UF followed by PEG precipitation allowed detection of 1000 cells of ST and SPA seeded in 20 L (0.05 cells per ml) of sewage. Application of this method in a low-resource neighborhood in Dhaka, Bangladesh indicated that ST was present in 26.7% (8/30) of open drain samples and 100% (4/4) of canal samples. Moore swabs, followed by enrichment culture, could detect as few as 100 seeded ST and SPA cells in 20 L (0.005 cells per ml) of sewage in laboratory experiments. Application of this detection method at three sewage pumping stations in Kolkata, India indicated that 69.8% of Moore swab samples were positive for ST and 20.8% were positive for SPA. These results demonstrate that UF and Moore swabs are two sensitive and effective methods for concentrating and detecting ST and SPA in wastewater and other environmental water samples, and these methods can be used for typhoid environmental surveillance and risk assessment.

In typhoid-endemic areas of Nepal and Bangladesh, previous investigators have reported the detection of ST in grab samples of drinking water that were concentrated by membrane filtration, followed by DNA extraction from the filter and PCR analyses.^21, 26^ Moore swabs were instrumental in the detection of ST in sewage canals near Santiago, Chile that were used to irrigate vegetable crops typically eaten without cooking.^6^ These findings indicate risk of ST transmission via drinking water and raw produce, but these samples and methods do not provide quantitative information that could be used to estimate the burden of typhoid infection in the population. Quantitative detection of ST and SPA in wastewater samples collected from sites where the population catchment can be estimated allows modeling approaches to estimate prevalence of typhoid and paratyphoid infection in the catchment population.^9^ This goal requires careful selection of sample collection sites and reliable methods to detect ST and SPA in samples that represent excreta from a defined population – typically wastewater samples. Ideally, methods to detect ST and SPA in wastewater for environmental surveillance are quantitative, sensitive, specific, low-cost, and feasible for laboratories in low-resource settings.

Currently, two types of samples have been used for ST and SPA wastewater surveillance: grab samples and trap samples using Moore swabs.^10^ Grab samples are typically concentrated through some type of filtration or centrifugation, sometimes allowed to incubate in an enrichment broth, and then analyzed by culture or PCR. Grab samples can be collected either in small volume or large volume, but microbial concentration from large volume samples can improve the sensitivity of detection. However, the volume of a grab sample that can be feasibly processed may be limited by the turbidity of the wastewater or environmental water that can clog filters used for sample concentration. Grab samples are usually analyzed without enrichment in order to obtain quantitative estimates of the pathogen concentration in the sample. In contrast, trap samples, such as Moore swabs, continuously filter and trap microbial pathogens in flowing water or wastewater and can be used successfully for turbid waters and wastewater. Moore swabs are often analyzed with primary enrichment, followed by PCR or culture methods, to increase the detection sensitivity. When an enrichment step is included in processing of grab samples or trap samples, the results only indicate the presence/absence of ST or SPA in the sample. One important difference between grab samples and trap samples is their ability to capture pathogen presence in wastewater in settings where there is large temporal variability. Results from grab samples reflect the presence/absence and concentration of a pathogen at a single point in time when the sample is collected. Moore swabs concentrate pathogens present in wastewater or environmental waters over a period of time (usually days), and therefore can be useful for sampling sites that represent smaller catchment populations where pathogen shedding may be infrequent and intermittent.

UF is an effective technique for concentrating multiple microbes simultaneously. Laboratory studies that seeded selected viruses, bacteria, and parasites into large-volume samples of tap water and reclaimed wastewater (100 L) reported mean recovery rates of five bacteria, protozoa, and viruses between 38 to 130%.^17, 27^ Two UF techniques, hollow-fiber ultrafiltration (TFUF) using tangential-flow^28^ and dead-end ultrafiltration (DEUF),^29^ provide effective recovery of diverse microbes from different types of water and wastewater, and both methods have showed similar recovery efficiencies in previous studies.^28, 29^ The main difference between the DEUF technique and TFUF configuration is that one of the ultrafilter ports is closed so that the water/wastewater sample must pass through the membrane. In contrast, the TFUF technique allows particles and other constituents larger than the membrane pore to be circulated and concentrated into a smaller volume. The DEUF technique is easier to perform than the TFUF because the water passes through the ultrafilter only once. However, the DEUF approach is more likely to clog the ultrafilter than the TFUF configuration and therefore is more appropriate for samples with low turbidity (e.g., tap water or reclaimed water). In this study, we used the hollow-fiber TFUF technique to concentrate turbid samples (e.g., from open drains and canals). With this method, particles and microbes are maintained in the ultrafilter cartridge under pressure, and there is a low risk of the ultrafilter clogging.^28^ Although the UF method has been described previously,^17, 27^ our recent deployment of this technique to process field samples in Dhaka, Bangladesh demonstrated the feasibility and value of this method to detect a range of pathogens, including *S*. Typhi, in high proportions of drain, canal, and flood water samples in a low-resource setting.^18^

“Moore swabs” have been used for decades by public health professionals around the world to detect and isolate enteric pathogens from wastewater and environmental waters, and their use to recover typhoidal *Salmonella* bacteria has recently been reviewed by Sikorski and Levine.^6^ Consisting of a strip of cotton gauze tied with string and suspended in flowing water, this sampling method acts as a trap that allows collection of pathogens over an extended period of time, especially when pathogen shedding is intermittent and in sites with smaller catchment populations. The Moore swab was first deployed by Brendan Moore in 1948 to trace *Salmonella* Paratyphi B from sewage in North Devon, England to determine the sources of contamination responsible for sporadic outbreaks of paratyphoid fever. Since the first application to detect typhoidal *Salmonella* in sewage, the Moore swab method has been utilized in several studies throughout the world to detect *Salmonella* Typhi in irrigation water, surface water, municipal sewers, and storm drains.^11, 30-32^ Given its effectiveness, simplicity, and affordability, we adapted this method for use in typhoid environmental surveillance of wastewater in Kolkata, India. We found that the Moore swab method has several advantages over UF in terms of greater sensitivity, simplicity, shorter processing time, less labor, and lower cost. However, the Moore swab method also has several limitations. First, this method only provides results on the presence or absence of ST and SPA in the sample and does not allow a quantitative assessment of ST or SPA concentration in the sample. Second, Moore swab sample collection requires two trips to place and later retrieve the swab, whereas grab samples only require a single collection trip. Third, deployment of Moore swabs for environmental surveillance has not been standardized in terms of sampling frequency, duration of immersion, and swab processing. This study addresses some of these information gaps by providing a standardized processing method for ST and SPA detection in wastewater with some benchmarks for limit of detection in wastewater matrices. We recognize that wastewater is a complex and highly variable matrix and that the limit of detection for this method will vary by setting.

The Moore swab method described in this study provides a low-cost, simple, and sensitive approach for presence/absence detection of ST and SPA in wastewater samples and is feasible to deploy in a wide range of low-resource settings. In addition, the UF method we have developed allows sensitive quantitative detection of ST and SPA in large volume (20-80 L) samples of wastewater. These two methods can be used in tandem or separately depending on the environmental setting and the surveillance or research purpose. Typhoid environmental surveillance strategies could start by deploying low-cost Moore swabs at a large number of sites in a screening stage to determine which locations provide the most valuable information. For sites where ST or SPA is detected in a Moore swab sample, follow-up large volume samples could be collected and processed by UF to obtain quantitative estimates of ST or SPA concentration which could be used to estimate infection prevalence in specific catchment populations. This type of two-stage approach for typhoid environmental surveillance would optimize use of human and financial resources and allow dynamic adaptive sampling site allocation that will more rapidly identify typhoid hotspots.^9^

## Data Availability

All data is available upon request

## ACKNOWLEDGEMENT

This study was funded by The Bill & Melinda Gates Foundation (BMGF) grant OPP1150697. We are in debt to Supriya Kumar and Megan Carey from the BMGF for their support and guidance. We are grateful to the environmental sample collection team from the National Institute of Cholera and Enteric Diseases (NICED), Kolkata, India and Infectious Disease Division, International Centre for Diarrhoeal Disease Research, Bangladesh (ICDDR,B) for their efforts to identify sewage sampling locations and their assistance with sample collection.

